# Revisiting the diagnostic criteria for emergence from the minimally conscious state: An empirical investigation

**DOI:** 10.1101/2021.11.30.21265893

**Authors:** Katherine Golden, Kimberly S. Erler, John Wong, Joseph T. Giacino, Yelena G. Bodien

## Abstract

**Objective:** To determine whether consistent command-following (CCF) should be added to the diagnostic criteria for emergence from the minimally conscious state (MCS)

**Design:** Retrospective cohort study

**Setting:** Inpatient rehabilitation hospital

**Participants:** Patients with severe acquired brain injury and disorders of consciousness (DoC) admitted to a specialized rehabilitation program

**Main Outcome Measure:** Difference between time to recovery of CCF and time to recovery of functional object use [FOU] or functional communication [FC] (the two existing criteria for emergence from MCS) as measured by the Coma Recovery Scale – Revised [CRS-R]).

**Results:** Of 214 patients (median [interquartile range] age: 53 [34, 66] years, male: 134 (62.6%), traumatic etiology: 115 (53.7%), admission CRS-R total score: 10 [7, 13]) admitted to rehabilitation without CCF, FO, or FC, 162 (75.7%) recovered CCF and FOU or FC during the eight-week observation period. On average, recovery of CCF, FOU, and FC was observed within one day of one another, approximately 46 [38.25, 58] days post-injury. One hundred and sixteen patients (71.6%) recovered FOU or FC prior to or at the same time as CCF.

**Conclusions:** In patients recovering from DoC, CCF reemerges around the same time as FOU and FC. This finding likely reflects the shared dependency of these behaviors on cognitive procecess (e.g., language comprehension, attention, motor control) that are essential for effective interpersonal interaction and social participation. Our results support the addition of CCF to the existing diagnostic criteria for emergence from MCS.

## Introduction

In 2002, the Aspen Neurobehavioral Conference Workgroup developed diagnostic criteria for the minimally conscious state (MCS) and emergence from MCS (eMCS)^1^: two key recovery milestones for patients with disorders of consciousness (DoC). MCS is characterized by clearly discernible, but fluctuating signs of consciousness (e.g., visual tracking and command-following). Recovery of functional object use (FOU) *or* functional communication (FC) (abbreviated FOU/FC below) signal eMCS.^1^ These behaviors were selected as eMCS criteria based on their role in supporting functional independence and effective interpersonal interactions. The diagnostic criteria for eMCS were established by expert consensus and have not undergone evidentiary validation.

The Coma Recovery Scale – Revised (CRS-R)^1,2^ is among the most psychometrically robust measures for establishing a DoC diagnosis^3^ and includes direct evaluation of behaviors consistent with MCS and eMCS. However, at least two studies suggest that even conscious patients with severe brain injury may have difficulty achieving the operational threshold for emergence from MCS.^4,5^ Moreover, the United Kingdom National Guidelines on DoC include visual discrimination, one approach for assessing command-following, among the eMCS diagnostic criteria.^6^ Behaviors other than FOU and FC, such as consistent command-following (CCF), may share a common neurobiological substrate and have similar cognitive processing demands.^7^ Identifying the full range of behaviors associated with reemergence of personal agency is consistent with the original intent of establishing criteria for eMCS and may help avoid delayed initiation of comprehensive inpatient rehabilitation services.

We tested the hypothesis that CCF recovers at approximately the same time as FOU/FC. We chose to study CCF based on the premise that expression of CCF and FOU/FC are dependent upon preserved connectivity of the language network and that these behaviors have similar reliance on linguistic decoding, vigilance, response persistence, and motor planning. Like FOU/FC, CCF is essential for active engagement in rehabilitation and is a strong independent predictor of functional recovery.^8^

## Methods

The Mass General Brigham Institutional Review Board provided ethical approval for this study. Written informed consent was not required because all data were obtained from the electronic medical record. Data were stored in REDCap (Research Electronic Data Capture), a secure, web-based data capture tool.^9^

### Measures

The CRS-R is a standardized neurobehavioral assessment instrument designed to evaluate auditory, visual, motor, oromotor, communication, and arousal functions in patients with DoC.^2^ Transition from MCS to eMCS is marked by the presence of either: 1) appropriate use of two different objects (FOU) *or* 2) accurate responses to six consecutive yes/no situational orientation questions (FC), on two consecutive CRS-R examinations. CCF is operationally defined on the CRS-R as four consecutive accurate responses to two different commands.

### Participants

We included patients who met the following criteria: 1) diagnosis of severe traumatic or non-traumatic acquired brain injury resulting in DoC; 2) at least 16 years of age; 3) admitted to a comprehensive inpatient rehabilitation program; 4) no evidence of CCF or FOU/FC on initial CRS-R administration; and 5) recovered FOU/FC and CCF during a pre-specified eight-week observation period. We excluded patients who: 1) were not admitted for inpatient rehabilitation immediately after discharge from acute care (n=5), 2) did not have any valid CRS-R examinations during the observation period (n=15 with no exams; n=8 with only invalid exams), 3) had evidence of CCF or FOU/FC on admission (n=55), or 4) did not recover CCF or FOU/FC during the observation period (n=108).

### Procedures

Trained clinicians administered the CRS-R twice weekly over the eight-week observation period. We divided patients into five groups: (1) Group1_CCF+FOU/FC_: recovered CCF *and* FOU/FC on the same day; (2) Group2_CCF→FOU/FC_: recovered CCF prior to FOU/FC; (3) Group3_FOU/FC→CCF_: recovered FOU/FC prior to CCF; (4) Group4_CCFonly_: recovered CCF without recovery of FOU/FC; and (5) Group5_FOU/FConly_: recovered FOU/FC without recovery of CCF. For each group, we evaluated the days between injury and the first instance of CCF and FOU/FC.

### Statistical Analysis

We report descriptive statistics for demographic and clinical characteristics, Kruskal-Wallis tests with Bonferroni correction and Chi-square for group comparisons, and Mann-Whitney tests to compare days from injury to recovery of behaviors. We conducted analyses in IBM SPSS v24. Statistical significance was concluded if p < 0.05.

## Results

Among 405 patients with DoC admitted for inpatient rehabilitation between 2012 and 2020, 214 (median [interquartile range] age: 53 [34, 66] years, male: 134 (62.6%), traumatic etiology: 115 (53.7%), admission CRS-R total score: 10 [7, 13]) met inclusion criteria. Demographics did not differ between groups (see Table). Admission CRS-R total scores differed between groups (p<.001) and were lowest in Group4_CCFonly._

**Table.** Study Sample Characteristics.

**Table. Study Sample Characteristics**
|  | Sample<br>N=214 | Group 1<br>CCF+FOU/FC<br>N=60<br>(28%) | Group 2<br>CCF→FOU/FC<br>N=46<br>(21.5%) | Group 3<br>FOU/FC→CCF<br>N=56<br>(26.2%) | Group 4<br>CCF only<br>N=21<br>(9.8%) | Group 5<br>FOU/FC only<br>N=31<br>(14.5%) | P |
| --- | --- | --- | --- | --- | --- | --- | --- |
| Age (years) | 53<br>[34, 66] | 52<br>[37.75, 60.5] | 58<br>[29.5, 69.75] | 53<br>[28.5, 62.25] | 59<br>[47, 66] | 50<br>[31.5, 67.5] | <sup>a</sup>.515 |
| Sex-Male<br>N (%) | 134<br>(62%) | 37<br>(61%) | 34<br>(73%) | 34<br>(60%) | 13<br>(61%) | 16<br>(51%) | <sup>b</sup>.376 |
| Etiology-TBI<br>N (%) | 115<br>(53%) | 33<br>(55%) | 21<br>(45%) | 38<br>(67%) | 7<br>(33%) | 16<br>(51%) | <sup>b</sup>.054 |
| Injury - Admit<br>(days) | 26<br>[21, 33] | 26<br>[20, 33] | 26.5<br>[22, 30.75] | 26.5<br>[20.75, 34.5] | 24<br>[20, 41] | 26<br>[22.5, 33] | <sup>a</sup>.962 |
| Admission<br>Diagnosis (%)<br>VS/MCS-/MCS+ | 25/48/26 | 18/56/25 | 43/30/26 | 14/51/33 | 52/33/14 | 12/64/22 | <sup>b</sup>.001 |
| Admission<br>CRS-R<br>Total Score | 10<br>[7, 13] | 11<br>[8.75, 14] | 8<br>[4, 11.75] | 10<br>[7.75, 13.25] | 6<br>[4, 9] | 11<br>[8, 13] | <.001<sup>a</sup> |
| First Observed<br>FOU, FC or<br>Both (%) | 86/65/42 | 36/28/35 | 35/41/24 | 43/41/16 | NA | 77/19/3 | <.001<sup>b</sup> |

Data are median (IQR) unless indicated. Group1<sub>CCF+FOU/FC</sub>, Group2<sub>CCF→FOU/FC</sub> and Group3<sub>FOU/FC→CCF</sub> recovered CCF and FOU/FC within eight weeks of rehabilitation admission. After Bonferroni correction, admission CRS-R total scores differed between Group1<sub>CCF+FOU/FC</sub> and Group2<sub>CCF→FOU/FC</sub>, Group1<sub>CCF+FOU/FC</sub> and Group4<sub>CCFonly</sub>, Group3<sub>FOU/FC→CCF</sub> and Group4<sub>CCFonly</sub>, and Group4<sub>CCFonly</sub> and Group5<sub>FOU/FConly</sub>. Group5<sub>FOU/FConly</sub> was more likely to demonstrate FOU rather than FC as the first sign of eMCS.
Abbreviation: CCF *consistent command following*; CRS-R *Coma Recovery Scale – Revised*; eMCS *emergence from the minimally conscious state*; FC *functional communication*; FOU *functional object use*; MCS- *minimally conscious state (MCS) without evidence of language function*; MCS+ *MCS with evidence of language function*; TBI *traumatic brain injury*; VS *vegetative state*.
Statistical tests: <sup>a</sup>= Kruskal-Wallis Test; <sup>b</sup>= Pearson's Chi-squared Test

For patients in Group1_CCF+FOU/FC,_ Group2_CCF→FOU/FC_ and Group3_FOU/FC→CCF_ (n=162), days from injury to recovery of CCF and FOU/FC did not differ (injury to CCF = 46.5 [39.25, 59] days; injury to FOU/FC = 46 [38.25, 58] days, p=0.563, see Figure). Across patients in Group1_CCF+FOU/FC,_ Group2_CCF→FOU/FC_, and Group3_FOU/FC→CCF_, 71.6% recovered CCF either concurrently with or after FOU/FC. Group4_CCFonly_ (n=21) recovered CCF 71 [54, 86] days following injury. Group5_FOU/FConly_ (n=31) recovered FOU/FC 46 [39.5, 62] days after injury. In all groups except Group5_FOU/FConly_, eMCS was as likely to be diagnosed based on the presence of FOU as it was based on FC. In Group5_FOU/FConly_, patients were more likely to meet the diagnostic criteria for eMCS based on the presence of FOU (p<.001).

**Figure.**
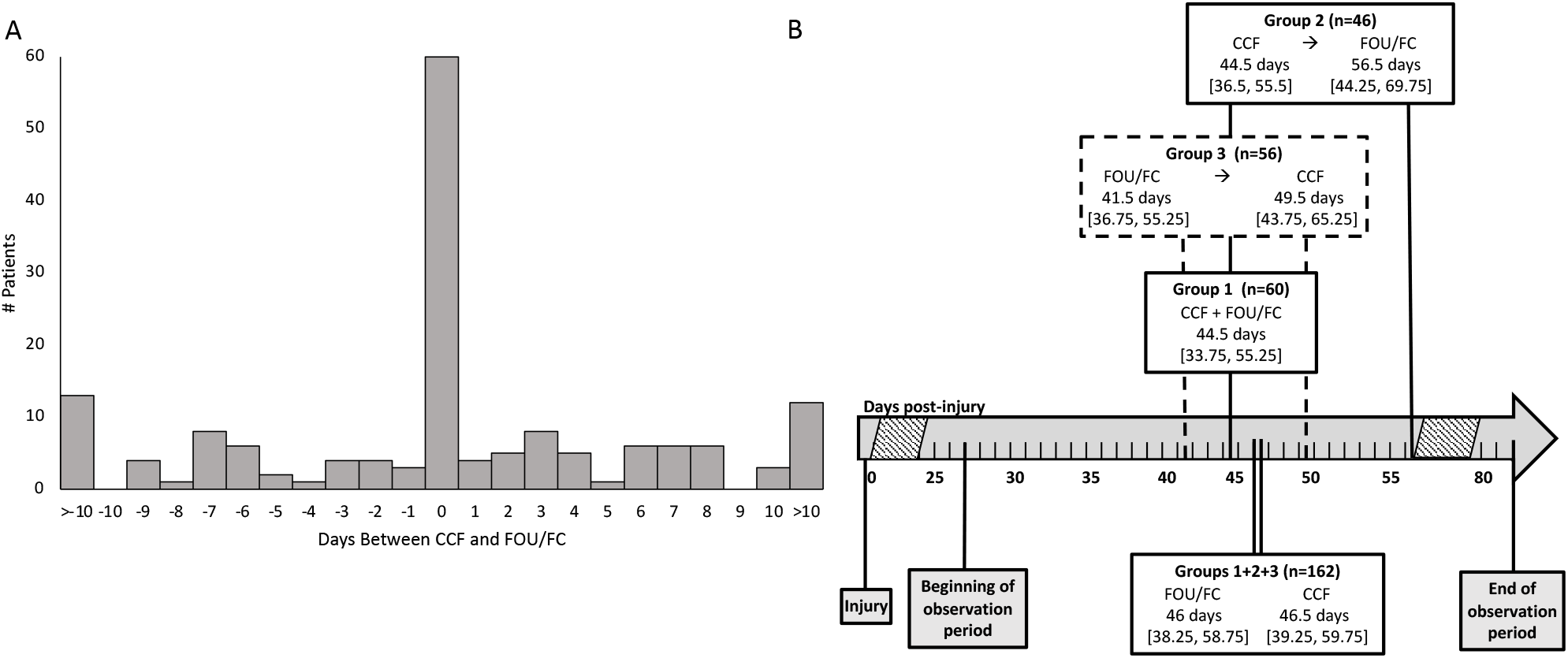
**A**. Temporal association between recovery of CCF and FOU/FC. On the x-axis, day 0 represents day of recovery of FOU/FC. Sixty patients recovered CCF on the same day as FOU/FC. Patients left of “Day 0” recovered CCF before FOU/FC; patients to the right of “Day 0”, recovered CCF after eMCS. **B**. Timeline of recovery of CCF and eMCS (i.e., FOU/FC). Days are reported using medians. During the observation period, Group1_CCF+FOU/FC_ recovered CCF and FOU/FC on same assessment; Group2_CCF→FOU/FC_ recovered CCF before FOU/FC; Group3_FOU/FC→CCF_ recovered FOU/FC before CCF. Abbreviation: CCF *consistent command following*; CRS-R *Coma Recovery Scale – Revised;* eMCS *emergence from the minimally conscious state*; FC *functional communication*; FOU *functional object use*; TBI *traumatic brain injury*.

## Discussion

To determine whether the criteria for eMCS should be extended to include CCF, we assessed the temporal relationship between recovery of CCF and FOU/FC. Consistent with a prior study reporting that CCF and FOU/FC frequently co-occur^7^, we found that CCF recovered at approximately the same time as FOU/FC. We reasoned that CCF is similar to FOU/FC because these behaviors rely on similar cognitive processes (e.g. language comprehension, attentional control, motor planning), which support return of effective communication and active participation in rehabilitation.

Approximately 10% of our sample recovered CCF but not FOU/FC (Group4_CCFonly_) during the observation period. These patients had the lowest admission CRS-R scores, suggesting a slower recovery trajectory. Approximately 15% of our sample recovered FOU/FC but not CCF (Group5_FOU/FConly_). This was the only group in which most patients (77%) demonstrated FOU as the first sign of eMCS, possibly reflecting impairment in language function.

### Study Limitations

Our sample is comprised of patients admitted to a specialized inpatient rehabilitation program for DoC and may not generalize to other settings. In addition, daily, rather than twice weekly, CRS-R assessment may have revealed a more precise trajectory for recovery of CCF and FOU/FC. Similarly, a longer observation period may have identified more patients who recovered all three behaviors and alternate recovery patterns. Finally, the immediate and long-term functional correlates of recovering CCF and FOU/FC across different time-scales are unknown. Although these behaviors recover together, their impact on subsequent recovery of independence requires external validation. Other approaches to data analysis, such as Item Response Theory, could provide further empirical support for including CCF as a criterion for eMCS.

### Conclusion

Recovery of CCF appears to follow the same trajectory as FOU/FC, suggesting these three behaviors may share similar mechanisms of action and have similar processing demands. These findings suggest that the diagnostic criteria for eMCS should be extended to include CCF. This modification of the existing diagnostic criteria for eMCS may facilitate early detection of this condition, improve individualized treatment planning, facilitate prognostication and help avoid delayed initiation of comprehensive inpatient rehabilitation services in this population.^10^

## Data Availability

All data produced in the present study are available upon reasonable request to the authors

## Acknowledgements

The contents of this publication were developed under grants from the National Institute on Disability, Independent Living, and Rehabilitation Research (90DPTB0011), which is a Center within the Administration for Community Living (ACL), Department of Health and Human Services (HHS), Tiny Blue Dot Foundation and the James S. McDonnell Foundation. The authors declare no relevant conflict of interest.

## Abbreviations

CCF: Consistent Command-Following
CRS-R: Coma Recovery Scale – Revised
DoC: Disorders of Consciousness
eMCS: Emergence from Minimally Conscious State
FC: Functional Communication
FOU: Functional Object Use I
QR: Interquartile Range
MCS: Minimally Conscious State

## Notes

### Competing Interest Statement

The authors have declared no competing interest.

### Author Declarations

The Mass General Brigham Institutional Review Board provided ethical approval for this study. Written informed consent was not required because all data were obtained from the electronic medical record.

